# Enisamium is an inhibitor of the SARS-CoV-2 RNA polymerase and shows improvement of recovery in COVID-19 patients in an interim analysis of a clinical trial

**DOI:** 10.1101/2021.01.05.21249237

**Authors:** Olha Holubovska, Denisa Bojkova, Stefano Elli, Marco Bechtel, David Boltz, Miguel Muzzio, Xinjian Peng, Frederico Sala, Cesare Cosentino, Alla Mironenko, Jens Milde, Yuriy Lebed, Holger Stammer, Andrew Goy, Marco Guerrini, Lutz Mueller, Jindrich Cinatl, Victor Margitich, Aartjan J. W. te Velthuis

## Abstract

Pandemic SARS-CoV-2 causes a mild to severe respiratory disease called Coronavirus Disease 2019 (COVID-19). Control of SARS-CoV-2 spread will depend on vaccine-induced or naturally acquired protective herd immunity. Until then, antiviral strategies are needed to manage COVID-19, but approved antiviral treatments, such as remdesivir, can only be delivered intravenously. Enisamium (laboratory code FAV00A, trade name Amizon®) is an orally active inhibitor of influenza A and B viruses in cell culture and clinically approved in countries of the Commonwealth of Independent States. Here we show that enisamium can inhibit SARS-CoV-2 infections in NHBE and Caco-2 cells. *In vitro*, the previously identified enisamium metabolite VR17-04 directly inhibits the activity of the SARS-CoV-2 RNA polymerase. Docking and molecular dynamics simulations suggest that VR17-04 prevents GTP and UTP incorporation. To confirm enisamium’s antiviral properties, we conducted a double-blind, randomized, placebo-controlled trial in adult, hospitalized COVID-19 patients, which needed medical care either with or without supplementary oxygen. Patients received either enisamium (500 mg per dose) or placebo for 7 days. A pre-planned interim analysis showed in the subgroup of patients needing supplementary oxygen (n = 77) in the enisamium group a mean recovery time of 11.1 days, compared to 13.9 days for the placebo group (log-rank test; p=0.0259). No significant difference was found for all patients (n = 373) or those only needing medical care (n = 296). These results thus suggest that enisamium is an inhibitor of SARS-CoV-2 RNA synthesis and that enisamium treatment shortens the time to recovery for COVID-19 patients needing oxygen.

**Significance statement:** SARS-CoV-2 is the causative agent of COVID-19. Although vaccines are now becoming available to prevent SARS-CoV-2 spread, the development of antivirals remains necessary for treating current COVID-19 patients and combating future coronavirus outbreaks. Here, we report that enisamium, which can be administered orally, can prevent SARS-CoV-2 replication and that its metabolite VR17-04 can inhibit the SARS-CoV-2 RNA polymerase *in vitro*. Moreover, we find that COVID-19 patients requiring supplementary oxygen, recover more quickly than patients treated with a placebo. Enisamium may therefore be an accessible treatment for COVID-19 patients.

## Introduction

Severe acute respiratory coronavirus 2 (SARS-CoV-2) is an important human pathogen and the causative agent of COVID-19. Vaccines are available to prevent the spread of SARS-CoV-2, and several antiviral strategies, such as treatment with remdesivir or reconvalescent plasma, have received emergency approval. However, the development of additional strategies remains necessary since vaccine roll-out is slow and current treatments can only be delivered intravenously. A key target for novel drug screening is the RNA polymerase of SARS-CoV-2 (1-4).

SARS-CoV-2 is a betacoronavirus and contains a positive-sense, non-segmented RNA genome of around 30 kilobases (5, 6). The 5’ two-thirds of the viral genome encode two overlapping open reading frames (ORFs), 1a and 1b, which are translated into two large polyproteins by host cell ribosomes. The two polyproteins are cleaved by intrinsic proteolytic activity to produce 16 non-structural proteins (nsps). Nsp12 is the RNA-dependent RNA polymerase that copies and transcribes the SARS-CoV-2 genome (7, 8). Nsp12 requires nsp7 and nsp8 for processivity *in vitro* (9) and likely other nsps, such as nsp9 and nsp13, for processivity *in vivo*. The structures of nsp12/7/8 and nsp8/9/12/13 complexes from SARS-CoV and SARS-CoV-2 have been solved by cryo-EM (10-12).

Remdesivir has been shown to inhibit the SARS-CoV-2 nsp12/7/8 complex and other nucleoside analogue drugs or small molecule inhibitors have been suggested as therapeutic candidates (1-3). One of the drugs highlighted by the World Health Organisation as a candidate therapeutic against SARS-CoV-2 is enisamium (4-(benzylcarbamoyl)-1-methylpyridinium) iodide (laboratory code FAV00A, marketed as Amizon®; Fig. 1A). Enisamium is licensed for use in 11 countries and a recent study found that enisamium is hydroxylated in humans and human lung cells to a compound called VR17-04 (Fig. 1A). VR17-04 inhibits the activity of the influenza virus RNA polymerase and reduces viral shedding and improves patient recovery in influenza patients (13).

**Figure 1.**
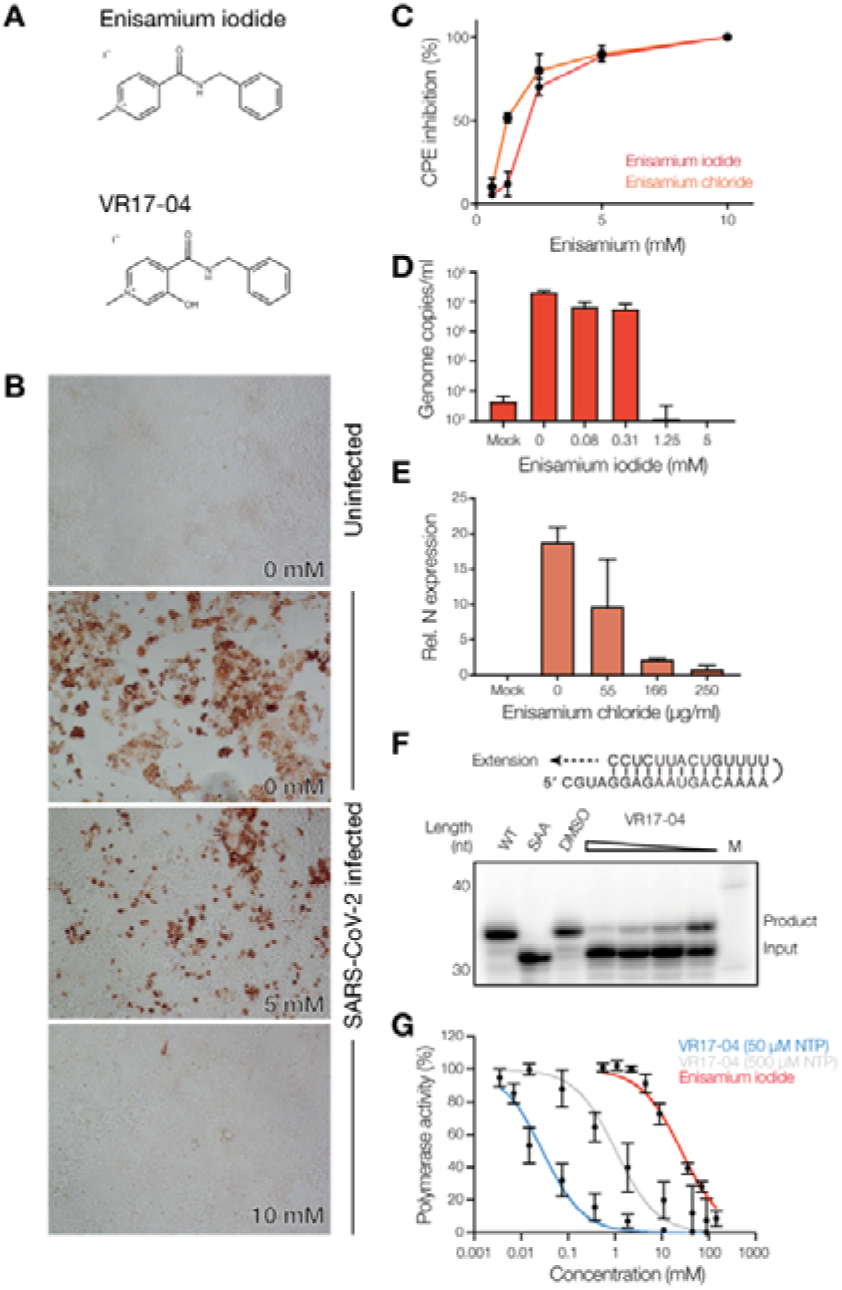
Enisamium inhibits SARS-CoV-2 infection and replication *in vitro*. (**A**) Chemical structures of FAV00A and VR17-04. The chemical structure of FAV00B is identical to FAV00A except that chloride ions are present instead of iodide. (**B**) Inhibition of SARS-CoV-2 N expression in Caco-2 cells by enisamium chloride. (**C**) Inhibition of SARS-CoV-2 cytopathic effect in Caco-2 cells by enisamium iodide and chloride. (**D**) Quantification of SARS-CoV-2 RNA genome levels in NHBE cells infected with SARS-CoV-2 after treatment with enisamium iodide. (**E**) Quantification of HCoV-NL63 N mRNA levels in NHBE cells infected with HCoV-NL63 after treatment with enisamium chloride. (**F**) Inhibition of the SARS-CoV-2 nsp12/7/8 RNA polymerase complex by VR17-04 on a hairpin template. A mutant containing a double amino acid substitution in the nsp12 active site (SDD=>SAA) was used as negative control. DMSO was used as solvent control. (**G**) Quantification of SARS-CoV-2 nsp12/7/8 RNA polymerase complex inhibition by enisamium or VR17-04 on a hairpin template. For VR17-04 two nucleotide triphosphate concentrations were used. Polymerase activity was plotted against drug concentration and dose-response curves were fit to the data. Quantification is from n=3 independently prepared reactions using the same nsp12/7/8 protein preparation. Error bars represent standard deviation.

In this study, we show that enisamium metabolite VR17-04 inhibits the SARS-CoV-2 nsp12/7/8 complex *in vitro*, likely by preventing incorporation of GTP and UTP. We also show that parent compound enisamium inhibits SARS-CoV-2 growth on Caco-2 and NHBE cells. In patients needing medical care and supplementary oxygen COVID-19 (Severity Rating 4 [SR 4] of the modified WHO ordinal scale), enisamium significantly improved the time to recovery compared to a placebo control. These data imply that enisamium is a possible therapeutic option for treating SARS-CoV-2 infection.

## Results

### Enisamium inhibits SARS-CoV-2 infection in cell culture

Previous experiments showed that enisamium (Fig. 1A) can efficiently inhibit influenza virus replication in normal human bronchial epithelial (NHBE) cultures and A549 cells, and to a lesser extent in Caco-2 cells (13). Previous experiments have also demonstrated that enisamium is not cytotoxic to these cells (13). To test if enisamium can inhibit pandemic SARS-CoV-2 replication in cell culture, we first incubated Caco-2 cells, a standard cell-line for SARS-CoV-2 infection *in vitro*, with enisamium iodide or enisamium chloride for 6 hours and subsequently infected the treated cells with SARS-CoV-2. After 48 hours, inhibition of viral infection was assessed by antigen staining for viral nucleoprotein expression and RT-qPCR for viral genome replication. We observed a significant reduction in both viral nucleoprotein expression (Fig. 1B), a reduction in the cytopathic effect of the SARS-CoV-2 infection on Caco-2 cells (Fig. 1C), and a 2-log reduction in the number of viral genome copies in the infected cells as a function of the enisamium concentration (Fig. S1). The IC_50_ for enisamium chloride in Caco-2 cells was 1.2 mM (∼300 µg/ml), which is comparable to the inhibitory effect of enisamium on influenza A virus replication in Caco-2 cells (13). To test if enisamium inhibited SARS-CoV-2 replication in NHBE cells, NHBE cells were incubated with enisamium iodide for 6 hours and subsequently infected with SARS-CoV-2. Analysis of the viral RNA levels in infected NHBE cells revealed an IC_50_ of 250 µg/ml (Fig. 1D). We obtained comparable results in NHBE cells infected with alpha-coronavirus HCoV NL63 (Fig. 1E), implying that enisamium can inhibit coronavirus replication.

### Enisamium and VR17-04 inhibit SARS-CoV-2 nsp12/7/8 activity

Previous experiments showed that enisamium and VR17-04 can inhibit the influenza A virus RNA polymerase *in vitro* (13). To determine if enisamium and its putative metabolite VR17-04 can inhibit the SARS-CoV-2 RNA polymerase, we used a SARS-CoV-2 RNA polymerase *in vitro* assay that involved nsp12 as the RNA-dependent RNA polymerase, and nsp7 and nsp8 as processivity factors. We expressed and purified SARS-CoV-2 nsp7, nsp8 and nsp12, and mixed them at a ratio of 2:2:1 to form a nsp12/7/8 complex. Next, we incubated the nsp12/7/8 complex with a hairpin template (Fig. 1F) in the presence of 0.5 mM of each nucleotide triphosphate (NTP) and varying concentrations of enisamium or the previously identified enisamium metabolite VR17-04. Enisamium inhibited nsp12/7/8 activity at relatively high concentrations, with an IC_50_ of 26.3 mM (Fig. 1G). By contrast, VR17-04 had an estimated IC_50_ of 0.98 mM on the hairpin template (Fig. 1G). These IC_50_ values are within an order of magnitude of those observed for the inhibition of the influenza virus RNA polymerase (13). Moreover, the IC_50_ value for VR17-04 on SARS-CoV-2 nsp12/7/8 is similar to remdesivir triphosphate in the presence of 0.5 mM NTPs in a comparable assay (14). A 10-fold reduction of the NTP concentration in the assay lowered the VR17-04 IC_50_ value to 0.029 mM (Fig. 1G), suggesting that VR17-04 is competing with NTP incorporation. Collectively, these data suggest that RNA synthesis by the SARS-CoV-2 nsp12/7/8 complex is inhibited by enisamium and its metabolite VR17-04 *in vitro*.

### Enisamium forms hydrogen bonds with adenine and cytosine in nsp12 and influenza polymerase active site

Previous studies (13, 15) and our data in Fig. 1A, suggest that enisamium acts through metabolite VR17-04 and inhibits the activity of the influenza A virus and SARS-CoV-2 nsp12 RNA polymerases. However, the mechanism of RNA synthesis inhibition by VR17-04 is not fully understood. We hypothesised that the additional OH-group of VR17-04 could support hydrogen bond formation with adenine and cytosine (Fig. 2A), creating two hydrogen bonds in total, while enisamium would form only one hydrogen bond with these bases. We expected that VR17-04 would not form hydrogen bonds with guanine or uridine (Fig. S2), suggesting that the inhibitory effect of VR17-04 would be dependent on the template sequence.

**Figure 2.**
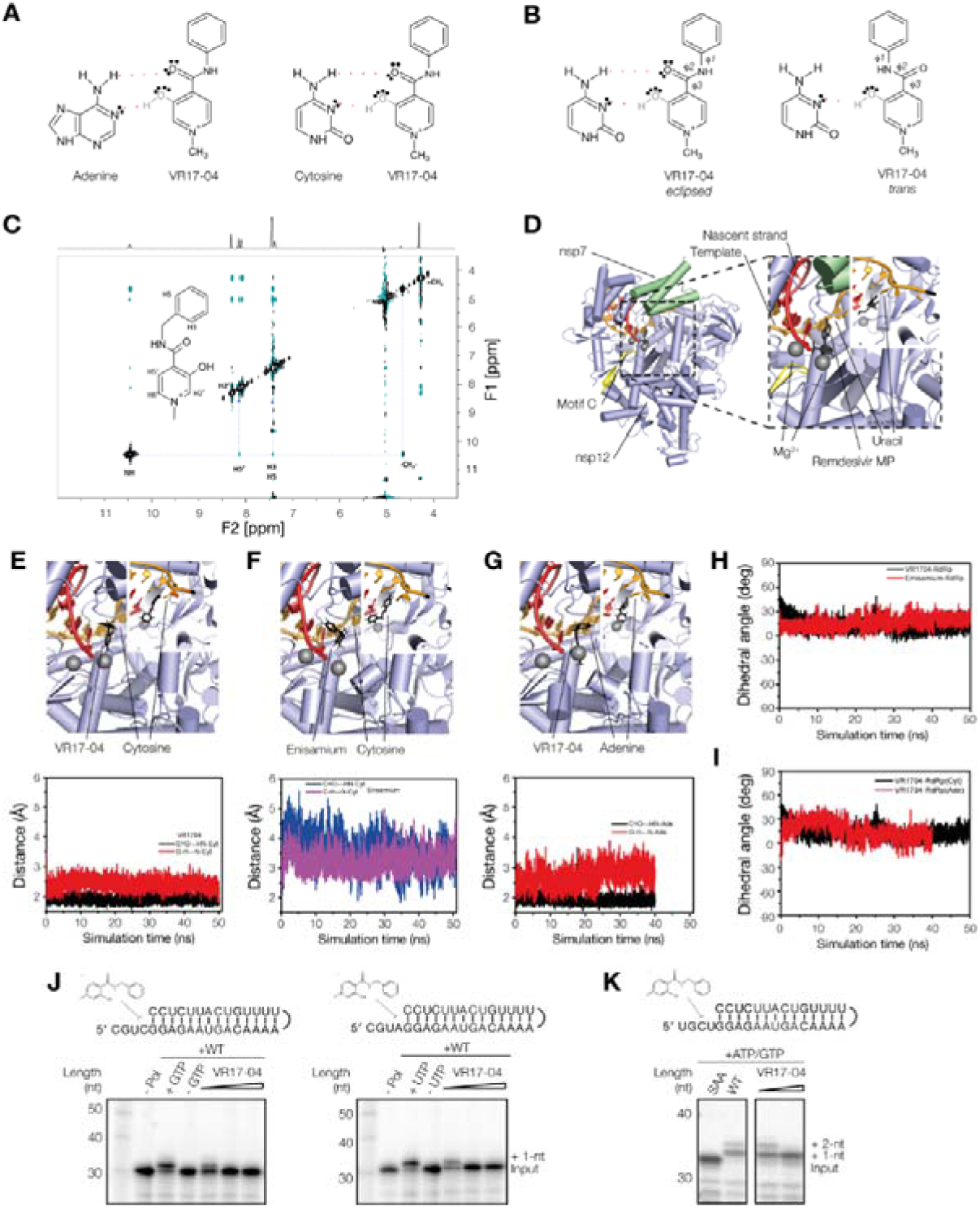
Enisamium metabolite VR17-04. (**A**) Schematic of putative hydrogen bond formation between cytosine and adenine bases with VR17-04. (**B**) Schematic of the *trans* and *eclipsed* conformations of VR17-04. (**C**) 2D-NOESY and 1H proton (above) spectra of VR17-04 acquired at 277 K in water. The NOE correlation between the HN and H5’ proton is highlighted with a dashed line. (**D**) Structure of the SARS-CoV-2 nsp12/7/8 complex bound to RNA and remdesivir monophosphate. Rendering based on PDB 7bv2. (**E**) Model (top) and MD simulation (bottom) of VR17-04 binding to cytosine in nsp12 active site. (**F**) Model (top) and MD simulation (bottom) of enisamium binding to cytosine in nsp12 active site. (**G**) Model (top) and MD simulation (bottom) of VR17-04 binding to adenine in nsp12 active site. (**H**) MD simulation of dihedral angle of VR17-04 or enisamium binding to cytosine in nsp12 active site. (**I**) MD simulation of dihedral angle of VR17-04 binding to cytosine or adenine in nsp12 active site (**J**) Effect of VR17-04 on SARS-CoV-2 nsp12/7/8 activity on two different hairpin templates in the presence of GTP (left) or UTP (right). In the presence of wildtype nsp12/7/8 and GTP or UTP, the radiolabelled primer was extended by 1 nt. (**K**) Effect of VR17-04 on SARS-CoV-2 nsp12/7/8 primer extension activity. A mutant containing a double amino acid substitution in the nsp12 active site (SDD=>SAA) was used as negative control. ATP and GTP were added to the reaction to allow extension of the template by 2 nt.

The CO and OH groups of VR17-04 can adopt *trans* and *eclipsed* conformations that are characterized by three dihedral angles: ϕ_1_, ϕ_2_ and ϕ_3_ (Fig. 2B). Dihedral angles ϕ_1_ and ϕ_2_ are also present in enisamium, but ϕ3 involves different atoms (Fig. S4). In VR17-04, dihedral angle ϕ_3_ is ∼180° in the *trans* conformation and ∼0° in the *eclipsed* conformation (Fig. 2B). Based on quantum chemical calculations, the *eclipsed* conformation has a lower energy than the *trans* transformation (Table S1), and only the *eclipsed* conformation would be compatible with cytosine or adenosine binding (Fig. 2B). To confirm that VR17-04 can adopt the *eclipsed* conformation in solution, we measured the ^1^H NOESY spectrum of VR17-04 in water and found a correlation between the protons ‘HN’ and ‘H5’ that is compatible with a ϕ3 value of 0° (Fig. 2C). The chemical shifts of the selected protons are reported in Table S2. Additionally, we observed a correlation between the HN and the CH_2_, or the ortho aromatic protons of the Ph group (Fig. 2C). These observations suggest that VR17-04 can adopt a conformation that would be compatible with hydrogen bond formation with cytosine and adenine.

To further investigate whether VR17-04 can inhibit the activity of the SARS-CoV nsp12 RNA polymerases through base-pair interactions with the template, we docked enisamium or VR17-04 into the SARS-CoV-2 nsp12/7/8 complex bound to template RNA and remdesivir monosphosphate (PDB 7bv2, Fig. 2D). Prior to docking, we removed the remdesivir monosphosphate from the complex, and used *in silico* mutagenesis to change the uridine in the active site to cytosine or adenine. After selection of the most significant poses, we found that both enisamium and VR17-04 can be accommodated in the +1 position of the nucleotide binding pocket (Fig. 2E-G), in a position similar to remdesivir monosphosphate (Fig. 2D). VR17-04 was specifically coordinated through hydrogen bond interactions with the unpaired cytosine residue of the template RNA. Nsp12 residues K545 and K555 were predicted to preserve the VR17-04 position in the catalytic cavity (Fig. S4). In addition, our modelling suggests that VR17-04 can form a stacking interaction with the -1 base of the nascent strand. By contrast, enisamium docked in the same +1 nascent strand position, but only formed one hydrogen bond with the cytosine in the +1 template position (Fig. S4).

To estimate the binding stability of enisamium or VR17-04 in the nsp12 active site, we performed molecular dynamics (MD) simulations of enisamium or VR17-04 in the nsp12 active site (see Material and Methods for specifics). Our MD simulations predict that VR17-04 favourably binds the unpaired cytosine in the +1 position of nsp12/7/8 complex, maintaining two hydrogen bonds (Watson-Crick base pair) as seen by the preserved hydrogen bonds distances (Fig. 2 E-G) and the coplanarity angle (Fig. 2H, I). By contrast, our simulations predict that enisamium binds less stably to the nsp12/7/8 complex, based on the ∼2-fold difference in distance between enisamium and cytosine compared to VR17-04 and cytosine (Fig. 2E, F; Table S3), and a higher estimated Poisson-Boltzmann free energy for the enisamium binding (43.6 kcal/mol) compared to VR17-04 binding to either cytosine (−19.8 kcal/mol) or adenine (−14.8 kcal/mol) (Table S4).

To investigate if VR17-04 inhibits the SARS-CoV nsp12/7/8 RNA polymerase complex in a sequence-specific manner, we analysed the extension of a hairpin template containing a cytosine or adenine in the +1 position of the template (Fig. 2J) in the presence or absence of VR17-04. We found that at high VR17-04 concentrations, the nsp12/7/8 RNA polymerase complex was prevented from incorporating GTP and UTP (Fig. 2J). When we subsequently moved the cytosine residue to the +2 nt position of the template down and introduced a uridine at the +1 position, ATP was incorporated in the presence of VR17-04 but GTP was not (Fig. 2K), suggesting that the inhibitory activity of VR17-04 is dependent on the sequence of the template.

### Enisamium Improves COVID-19 Patient Recovery in Interim Analysis of Clinical Trial

To investigate if enisamium affects the clinical course of COVID-19 patients, a double-blind, randomized, placebo-controlled phase III clinical study was performed. The COVID-19 diagnosis was based on a body temperature of ≥ 37.8 °C and laboratory confirmed presence of SARS-CoV-2 RNA by RT-PCR in pharyngeal swabs or sputum. The patient cohorts required either medical care but no supplementary oxygen (Severity Rating (SR) 5; WHO score 3), or medical care and supplementary oxygen (SR 4; WHO score 4) on the day of enrolment and randomization. Randomized COVID-19 patients were treated with either placebo or enisamium iodide for 7 days. The chosen primary endpoint of the trial was time-to-recovery, and recovery was defined as an improvement in the SR baseline status by 2 SR score values (e.g., a change from SR 4 to SR 6).

In accordance with the study protocol, an interim analysis on all patients in the Intent-to-Treat evaluation set (ITT) was performed for the primary endpoint by an Independent Data Monitoring Committee (IDMC) using a pre-defined Charter. The ITT set for interim analysis included 373 patients of which 296 patients had SR 5 and 77 patients had SR 4 on day of randomization (Fig. 3A). The interim analysis showed no difference in time-to-recovery and median time–to-recovery between all placebo- and enisamium-treated ITT patients (Fig. 3B) nor in the subgroup of patients with SR 5 (Fig 3C). In contrast, analysis of the SR 4 subgroup alone (n = 77) revealed a faster time-to-recovery with the enisamium-treated patients compared to the placebo-treated for the whole period of recovery, starting 7 days after randomization (Fig. 3D). The estimated median time–to-recovery was 13 days for the placebo-treated patients and 11 days for the enisamium-treated patients. The mean time-to-recovery was 13.9 days for the placebo-treated group and 11.1 days for the enisamium-treated group. The log-rank test showed a significant advantage in recovery time in favour of enisamium treatment at interim stage (*P* = 0.0259). The maximum time-to-recovery was reached on day 21 for the enisamium-treated group. Not all patients recovered in the placebo-treated group. For these patients, the recovery time was displayed as 29 days (Fig. 3D).

**Figure 3.**
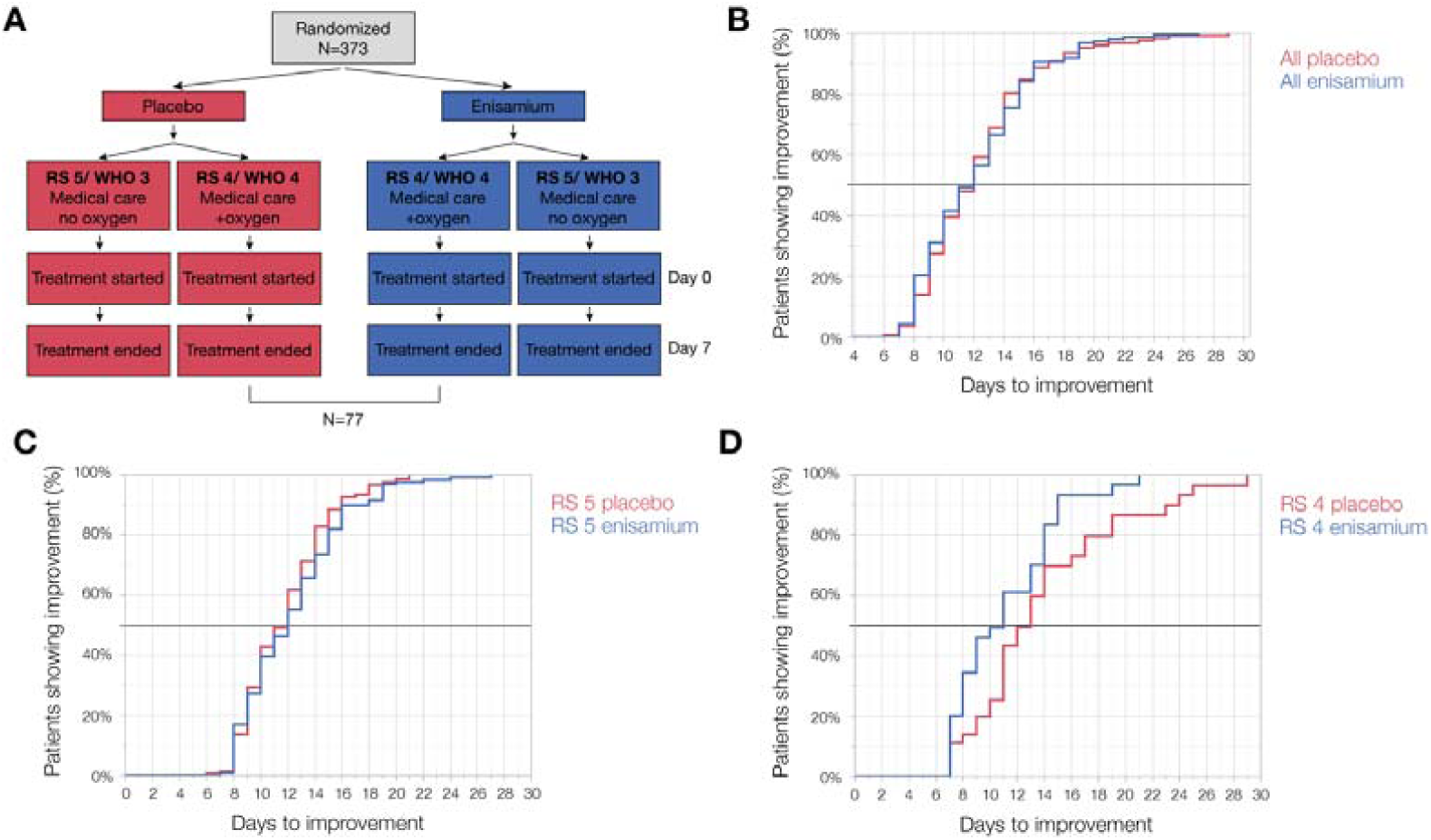
Enisamium improves recovery of COVID-19 patients requiring supplementary oxygen. (A) Schematic of patient recruitment, randomization and treatment. (B) Kaplan-Meier plot of the percentage improvement observed in all COVID-19 patients, (C) RS 5 COVID-19 patients (medical care with no oxygen support), and (D) RS 4 COVID-19 patients (medical care with non-invasive oxygen support).

Based on the above findings, the IDMC recommended to stop recruitment of patients requiring only medical care but no oxygen (SR 5; WHO score 3) and to continue the study with COVID-19 patients requiring supplementary oxygen in addition to standard medical care (SR 4; WHO score 4).

## Discussion

The rapid global spread of SARS-CoV-2 necessitates development of effective therapeutic interventions, and the most promising short-term strategy is to repurpose existing drugs. In this study we showed that enisamium, which is approved for use against influenza in 11 countries, can inhibit SARS-CoV-2 RNA synthesis (Fig. 1). Moreover, we showed that enisamium and its metabolite, VR17-04, inhibit the RNA synthesis activity of the SARS-CoV-2 nsp12/7/8 complex (Fig. 1, 2). Molecular dynamics analysis and *in vitro* activity assays suggest that VR17-04 prevents GTP and UTP incorporation into the nascent RNA chain (2).

It was previously reported that enisamium inhibits the influenza A virus RNA polymerase activity *in vitro* with a relatively high IC_50_ value of 46.3 mM (13). This inhibition was improved 55-fold by addition of a hydroxyl group in the compound VR17-04 (13). We find that SARS-CoV-2 nsp12/7/8 complex activity is inhibited by enisamium and VR17-04 with similar IC_50_ values to the influenza A virus RNA polymerase at similar NTP levels (Fig. 1) (13). Furthermore, the IC_50_ value for VR17-04 is similar to remdesivir triphosphate in a similar *in vitro* assay used (14). Remdesivir triphosphate is the active metabolite of remdesivir, which has shown promise in both cell culture and clinical trials as a treatment for SARS-CoV-2 infection (16).

Our docking and molecular dynamics simulations suggest that VR17-04 can bind a template cytosine or adenine base in the active site of the SARS-CoV-2 RNA polymerase. This hypothesis is supported by our NOE experiments performed in water, which indicate that VR17-04 can adopt an *eclipsed* conformation in solution. Only the *eclipsed* conformation is compatible with the sequence-specific inhibition we observed in our *in vitro* polymerase reactions. We cannot fully exclude the possibility that VR17-04 can also bind bases in the nascent RNA strand. However, we think that this alternative explanation does not adequately explain our data.

We observed that the inhibitory effect of enisamium was more pronounced in NHBE cells than in Caco-2 cells, which is in line with previous influenza A virus experiments (13, 15) and suggest that enisamium is more readily metabolised into VR17-04 in primary bronchial epithelial cells compared to adenocarcinoma cells. Interestingly, the interim phase III clinical trial results that we disclose here reveal a faster improved recovery in COVID-19 patients needing supplementary oxygen, strongly suggesting that our *in vitro* data are well-aligned with the data from the clinical trial. We cannot exclude that enisamium or its metabolite has effects beyond RNA polymerase inhibition and that these additional effects contribute to the COVID-19 patient recovery.

Overall, our results strongly suggest that enisamium metabolite VR17-04 inhibits RNA synthesis by the SARS-CoV-2 nsp12/7/8 complex. Together with the interim phase III clinical trial findings that enisamium improved the recovery of COVID-19 needing supplementary oxygen (SR 4) by more than 2 days, our observations raise the possibility that enisamium could be used a viable therapeutic option against SARS-CoV-2 infection. Moreover, unlike remdesivir, enisamium does not require intravenous administration, which would be advantages for its use outside of a hospital setting. Together with observations that enisamium can inhibit other RNA virus infections, and DNA virus infections (13, 15, 17), these results here suggest that it can act as broad-spectrum polymerase inhibitor.

## Materials and Methods

### SARS-CoV-2 infections

Confluent layers of Caco-2 cells in 96-well plates were treated with serial dilutions of FAV00A or FAV00B 6 hours prior to infection. The cells were infected with SARS-CoV-2 at multiplicity of infection 0.01 for 1 h, and compound reapplied following virus removal. At 48h post infection, the cytopathic effect was recorded by examination of infected cultures by light microscopy and supernatant collected to quantify virus RNA by RT-qPCR as described previously, using nsp12-specific primers 5’-GTGARATGGTCATGTGTGGCGG-3’ and 5’-CARATGTTAAASACACTATTAGCATA-3’ (18-20). Cells were fixed with acetone/methanol (40:60) and immunostained using a SARS-CoV-2 nucleoprotein monoclonal antibody (1:500, Sinobiological, Cat #40143-R019-100ul). Staining was detected using a peroxidase conjugated anti-rabbit secondary antibody (1:1000, Dianova) and the addition of AEC substrate.

### HCoV-NL63 NHBE infections

MatTek’s EpiAirway System (MatTek; Ashland, MA) consisted of differentiated NHBE cells that were cultured to form a multilayered, highly differentiated model that closely resembles the epithelial tissue of the respiratory tract. The cells from a single donor (No. 9831) were used for assay consistency. The apical surface of the cells was exposed to a humidified 95% air/ 5% CO2 environment. The basolateral medium was changed, and the mucin layer was washed every 24-48 hours. NHBE cells were inoculated via the exposure of the apical side to HCoV-NL63. After 1 hour of incubation with virus in a water-jacketed 37°C incubator with a 5% CO2 supply, the viral inoculum was removed from the cells. The apical side of the cells was washed once prior to infection. After viral inoculation, enisamium chloride or control media were added to the apical side of the cells and the basal media compartment and incubated with the cells for 1 hour. After a 1-hour incubation, the drug containing media was removed from apical and basal chambers. Growth medium alone or growth medium with enisamium chloride were added to the bottom chamber, and cells were incubated for 48 hours. At the termination of the experiments, cells were washed twice, then 1 ml Trizol (Invitrogen) was added to each well for RNA isolation. Total RNA was isolated from cells using Trizol per the manufacturer’s instruction. Two-step RT-qPCR was performed using HCoV-NL63 N gene-specific primers/probe (forward primer: 5’-TGGTGTTGTTTGGGTTGCTA-3’, reverse primer5’-GCTCTGGAGGCAAAGCAATA-3’, double-quenched probe: 5’-FAM/CGCAAACGT/ZEN/AATCAGAAACCTTTGGA/IABKFQ-3’), GAPDH was analysed at the same time with GAPDH-specific primers (5’-GTTCGACAGTCAGCCGCATC-3’ and 5’-AGTTAAAAGCAGCCCTGGTGA-3’) by RT-qPCR and served as a reference gene for normalization.

### SARS-CoV-2 nsp12/7/8 in vitro activity assays

Plasmids expressing nsp7, nsp8 and nsp12 were kindly provided by Dr Ervin Fodor (University of Oxford). Nsp7, nsp8 and nsp12 were purified as described previously (14) and mixed at a molar ratio of 2:2:1 to form active nsp12/7/8 complex. For activity assays, 50 nM hairpin template (5’-CGUAGGAGAAUGACAAAAUUUUGUCAUUCUCC-3’), or a variation of this template (see figures), was incubated with 1 µM nsp12/7/8 complex for 30 mins at 30 °C, in reaction buffer containing 5 mM MgCl_2_, 20 mM Hepes pH 8.0, 0.5 mM ATP, 0.5 mM UTP, 0.5 mM GTP, 0.5 mM CTP, 0.05% NP-40, 5% glycerol, 50 mM NaCl, 1 U RNasin (Promega) and 1 mM DTT. Hairpin templates were based on the 3’ terminal sequence of the SARS-CoV-2 genome and reference (3). Reactions were stopped by addition of 80% formamide and 10 mM EDTA, followed by heating to 95 °C for 3 min. Reaction products were resolved by 20 % denaturing PAGE with 7M urea, and visualised by phosphorimaging on a Typhoon FLA 7000 (GE Healthcare) scanner. Data were analysed using ImageJ and Prism 9 (GraphPad). Enisamium iodide (Farmak) or VR17-04 (Farmak) were dissolved in DMSO to a stock concentration of 250 and 125 mg/ml, respectively. Dilutions were made in DMSO.

### NOE NMR spectra measurement

The VR17-04 (2 mg) was dissolved in 0.6 ml D2O/water solution (5% D2O). NMR experiments were performed at 500MHz at 277 K. For NOESY experiments (noesygpph) 128 transients were collected for each free-induction decay, using a mixing time of 300 ms and 20 sec of relaxation delay. NOESY experiment (matrix 1024 · 320 points) was zero-filled to 2K · 2K before Fourier transformation. Measurements were taken on a Bruker 500 HD NMR spectrometer equipped with a 5 mm BB probe.

### Conformational characterization and geometry optimization

The VR17-04 and enisamium conformational characterization and geometry optimization was done using the quantum chemical approach DFT B3LYP/6-31G*. The lowest energy conformation of VR17-04 (ϕ_3_ *eclipsed*) and enisamium were selected for further docking and for partial charge estimation (Table S1).

### Molecular docking

The docking simulation was performed using Autodock 4.2 software (21). The geometry of the ligands was previously optimized by DFT B3LYP/6-31G*. The geometry of the receptor RdRp was extracted from the PDB 7bv2 (22). The catalytic site of the nsp12/7/8 complex include the template RNA strand, the nascent strand RNA, the pyrophosphate moiety [O_3_P-O-PO_3_]^-4^ (pyr) and two Mg^+2^ ions, whose positions were included in 7BV2. The two Zn^+2^ ions that were co-crystallized in 7BV2 were also included in the models. The co-crystallized inhibitor remdesivir was removed from the complex, while its position and the contacts with the template RNA trough the unpaired uracil base in the +1 position were used to guide the molecular docking. The unpaired uracil base was subsequently mutated in cytosine (Cyt) or adenine (Ade) in Pymol 2.3.4 (Schrodinger LLC), generating two different complexes, identified as nsp12/7/8(Cyt) and nsp12/7/8(Ade) respectively. Next, Gasteiger charges were calculated for both ligands (VR17-04, enisamium) and the receptor complexes, and used as parameters of the docking simulation (23). In the docking simulation VR17-04 and enisamium were described by five and four rotational degrees of freedom, respectively. The docking gridbox was built by orthogonal hedges of length between 60 to 80 points. The gridbox centre was set to the NH_2_-group of the target cytosine or adenine residues of the template RNA strand, respectively, and further set-up to fit the space between the R555 and K545, and the uracil of the nascent RNA strand. The docking runs used the default Genetic Algorithm search, with parameters: number of GA runs, population size, max number of energy evaluation, and max number of generations set as 100, 2000, 2.5.10^7^, 270.000. At each run the docking solutions were clustered using a tolerance RMSD = 2.0 Å. Three different docking simulations were run: VR17-04 and enisamium were docked on to nsp12/7/8(Cyt), while, for comparison, VR17-04 was further docked on nsp12/7/8(Ade). The docking solutions were selected based on two criteria. The first criterium was the possibility to form a Watson-Crick base-pair interaction between VR17-04 (or enisamium) and the unpaired cytosine or adenine. The second criterium was based on the possibility to figure out interferences of the ligand with the catalytic mechanism, for example by interaction with key residues of the nsp12/7/8 complex. The selected poses were further ranked by preliminary MD simulation (approximately 20 ns) in explicit solvent, to predict the stability of the interaction, and to estimate the Poisson-Boltzmann free energy of binding, this last property was used to obtain a final selection and ranking of the poses.

### Molecular dynamic simulation

Explicit solvent MD simulation were run using NAMD 2.12 (24) software, applying the Amber force field (ff14SB) (25). The TIP3P (26) water solvent model was used. The t-leap application of the Ambertools 14.0 package (25) was applied to generate the topology, the parameters, and the coordinate files of the macromolecular complex simulated. The coordinates of the macromolecular elements of the nsp12/7/8 complex: nsp12, nsp8, nsp7, pyr, template RNA, nascent strand RNA, were extracted from the PDB 7bv2. The geometries of the VR17-04, enisamium (before docking) and pyr were optimized using the quantum chemical approach DFT B3LYP/6-31G*; the corresponding partial charges were estimated fitting the electrostatic potential that was calculated at level of theory B3LYP/6-31G*/RHF/6-31G*. This procedure is in accord to the standard required by the Amber force field. The quantum chemistry software GAMESS (27) was used for this stage. The Amber atomtypes (parm10.dat) were selected for VR17-04, enisamium and pyr (antechamber application, Ambertools 14.0). Three macromolecular complexes indicated as VR1704-nsp12/7/8(Cyt), enisamium-nsp12/7/8(Cyt), and VR1704-nsp12/7/8(Ade), were solvated by a 15 Å wide layer of TIP3P water molecules in each X, Y, Z direction; the orthogonal simulation box was built with hedges of approximate length 116, 116, 127 Å. The non-bond electrostatic and dispersive interactions were described by the standard cut-off technique (12.0 Å). Before running MD simulation, each simulation cell box was energy minimized running 200 K steps of the default energy minimized algorithm, as implemented in NAMD. The MD simulations were run fixing the number of particle (N), the absolute temperature (T), and the pressure (P) applied to the cell hedges. The absolute temperature was 300 K and maintained with a Lowe–Andersen thermostat, while the pressure on the cell box hedges was set as P = 1.01325 bar and preserved by the Nosé–Hoover–

Langevin piston algorithm. The first MD simulation stage was run to adjust the simulation cell box density, allowing the relaxation of all the inter-molecule distances, i.e., the solute-solute, solute-solvent, and solvent-solvent distances. The cell density equilibration stage was run restraining the atoms of the solute to their initial position (energy minimized geometry of the complex) applying a harmonic restraint. In this stage of cell density equilibration, the water molecules were left free to move. The harmonic restraint constant value was set initially as 1 Kcal mol^-1^ for each atom of the solute, and progressively reduced at 0.5 and 0.2 Kcal mol^-1^. The cell density equilibration stage was monitored plotting the cell volume (Å^3^) *vs* time (ns), until the cell volume fluctuations level-off to a horizontal axis that corresponds to the average final volume of the cell. This stage required between 20 to 50 ns approximately and was further checked by the formation of a thin water shell that surround the face of the inhibitor molecule that is exposed to the empty catalytic cavity of the nsp12/7/8 complex. In the second stage of the MD simulation, the harmonic restraint was removed, and the inhibitor-nsp12/7/8 complex was allowed to equilibrate in the geometry and the relative position of their elements: inhibitor, template RNA, nascent strand RNA, pyr, Mg^+2^, and Zn^+2^. To monitor the equilibration of position and orientation that VR17-04 (or enisamium) presents in the +1 position of the catalytic cavity, the distances between the carbonyl oxygen of VR17-04 and the hydrogen (NH_2_-) of cytosine (or adenine), and between the hydroxyl group of VR17-04 and the nearby lone pair of cytosine nitrogen (or adenine), were plotted *vs* simulation time. These distances provide a direct indication of the stability of the Watson-Crick base pair interaction that hold the inhibitor and the target base. The orientation of the VR17-04 in the catalytic site of the nsp12 were also monitored by the improper dihedral angle ϕ defined by the following atoms (bold) C**O**-**N**H_2_-**N**-**O**H, in which C**O** and **O**H belong to VR17-04, while the remaining **N**H_2_ and **N** belong to the opposite cytosine (or adenine). Alternatively, the enisamium-cytosine pair require the following atoms C**O**-**N**H_2_-**N**-**C**H to define ϕ, in this case C**O** and **C**H belong to enisamium, while **N**H_2_ and **N** belong to the opposite cytosine. In fact, values of this angle around ‘0’ indicate that the ‘base pair’ contact between VR1704 the cytosine (or adenine) is coplanar, a geometric condition favouring the Watson-Crick ‘base-pair’ interaction between VR17-04 and the base. VMD 1.9.3 (28) was used for the MD simulation trajectory visualization and image creation.

### Estimation of Poisson-Boltzmann free energy of binding

In a system that evolve in accord to the complex formation reaction (29).

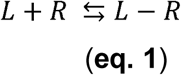

The free energy change is calculated knowing only the initial and final state of the system:

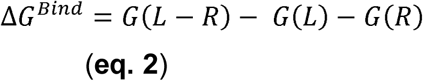

The free energy decomposed in the following terms:

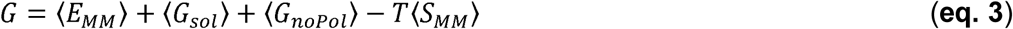

The free energy change is then conveniently split:

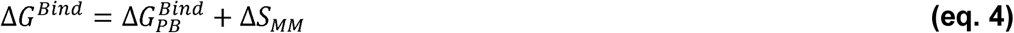

In **eq. 3** the *E*_*MM*_ correspond to the potential energy of the system, as described by the force-field; *G*_*sol*_ is the polar solvation energy, estimated by the Poisson-Boltzmann equation (30); *G*_*nopol*_ is the non-polar solvation energy, estimated by the solvent-accessible surface area, a method included in the MMPBSA. *T* is the absolute equilibrium temperature, while *S*_*MM*_ is the molecular entropy of the system. The sum of the first three terms on the right-hand side of the **eq. 3** is conveniently defined Poisson-Boltzmann free energy of binding *ΔG*_*PB*_^*Bind*^ (see **eq. 4**). To estimate the absolute value of the free energy of binding *ΔG*^*Bind*^ the molecular entropy change Δ*S*_*MM*_ is required. Since in this study two similar molecules: VR17-04 and enisamium were compared as bound to cytosine (+1 position of the catalytic site of nsp12), or alternatively two bound states VR17-04-cytosine or VR17-04-adenine were inquired, in a level zero approximation the respective entropy changes Δ*S*_*MM*_ are considered similar. In this condition the Poisson-Boltzmann free energy of binding *ΔG*_*PB*_^*Bind*^ could be used to rank the selected molecular recognitions and/or the poses of the docking experiment.

### Patients

For this prospective, multi-centre, randomized, double-blind, placebo-controlled, parallel-group phase III clinical trial, male and female hospitalized patients ≥ 18 years with moderate severity of COVID-19 infection were eligible for participation. The diagnosis of COVID-19 was based on body temperatures of ≥ 37.8 °C and laboratory confirmed RT-PCR SARS-CoV-2 test from pharyngeal swabs or sputum. According to intent-to-treat (ITT) definition and the actual recruitment status, 373 patients from 15 Ukrainian hospitals could be included into the interim analysis.

The study was conducted in accordance with the Declaration of Helsinki, ICH-GCP and the national laws and regulations in Ukraine. All patients singed an informed consent prior to study participation. The study was approved by the Ethics Commission of the Regional Clinical Hospital of Ivano-Frankivsk Regional Council on 12.05.2020. The registration number of the study in ClinicalTrials.gov is NCT04682873.

### Treatment

Patients were randomized at a 1:1 ratio to receive either 500 mg enisamium iodide or matching placebo 4 times daily every 6 h for 7 full days. Supporting treatment could be given at the investigator’s discretion. All patients treated with either placebo or enisamium, having a laboratory-confirmed SARS-CoV-2 at randomization and providing any valid efficacy data after initiation of treatment were included in the Intent-to-Treat evaluation set (ITT).

### Data Collection

Baseline clinical data of the patients were recorded in the electronic case report forms including demographics, medical history, previous and ongoing medication, body height and weight and results of physical examination. During the study up to day 29 or until discharge, symptom severity rating, vital signs, assessment of COVID-19 symptoms, safety lab results, concomitant medication and adverse events were recorded. The patient’s and investigator’s judgment on safety and efficacy were collected independently. RT-PCR tests on SARS-CoV-2 at defined intervals and at the discretion of the investigator were carried out.

### Primary Outcome

The primary outcome was measured as the time from the day of randomization (day 1) to an improvement of at least two score points (from the status at randomization) on the severity rating (SR) scale in days. The SR scale (WHO scale) is defined as 1 (8) - Death; 2 (7, 6) - Hospitalized, on invasive mechanical ventilation or extracorporeal membrane oxygenation; 3 (5) - Hospitalized, on non-invasive ventilation or high flow oxygen; 4 (5) - Hospitalized, requiring supplemental oxygen; 5 (3) - Hospitalized, not requiring supplemental oxygen – requiring ongoing medical care; 6 (−) - Hospitalized, not requiring supplemental oxygen – no longer requires ongoing medical care; 7 (2) - Not hospitalized, limitation on activities and/or requiring home oxygen; 8 (1) - Not hospitalized, no limitations on activities. Safety of enisamium iodide will be analysed based on the incidence of adverse events, vital signs, judgement of global tolerability on day of hospital discharge (separately by patient and investigator) and laboratory data when the study is completed.

### Statistical Analysis of Interim Data

The unblinded interim analysis was carried out by an Independent Data Monitoring Committee based on a pre-specified Charter. At this stage, the primary outcome was pre-planned to be tested by using the two-sided log-rank test stratified by centre. Because of the low patient number in some centres the stratification was omitted for gaining primary interim results. The primary outcome was defined as an improvement from baseline of at least two SR points and was evaluated for all ITT patients at the interim stage as well as for subgroups with baseline score of SR 4 and 5 separately. The interim analysis used the promising zone approach according to (31) that was implemented according to (32). For this method no adjustment of the type-I error rate is needed.

## Supporting information

Consort checklist

## Data Availability

The data will be made available on ClinicalTrials.gov via number NCT04682873 and via the provided supplemental information.

## Acknowledgments

The authors would like to thank Dr. Juergen Richt (Kansas State University, Manhattan, KS, US) for comments and advice, and Dr Ervin Fodor, Dr Jonathan Grimes, Alexander Walker, Dr Haitian Fan and Dr Jeremy Keown (University of Oxford, Oxford, UK) for expression constructs and sharing preliminary data.

## Funding

A.J.W.t.V is supported by joint Wellcome Trust and Royal Society grant 206579/Z/17/Z and the National Institutes of Health grant R21AI147172. Part of this research was funded by Farmak Public Joint Stock Company, Kiev, Ukraine.

## Conflict of interest

V.M. and A.G. are employees of Farmak Public Joint Stock Company, Kiev, Ukraine. Part of this research was funded by Farmak Public Joint Stock Company, Kiev, Ukraine.

## Notes

### Competing Interest Statement

V.M. and A.G. are employees of Farmak Public Joint Stock Company, Kiev, Ukraine. Part of the research was funded by Farmak Public Joint Stock Company, Kiev, Ukraine.

### Clinical Trial

NCT04682873

### Author Declarations

The study was approved by the Ethics Commission of the Regional Clinical Hospital of Ivano-Frankivsk Regional Council on 12.05.2020. The registration number of the study in ClinicalTrials.gov is NCT04682873.

### Summary of Updates

The legend in Fig. 3 was revised and the group A/B annotations replaced with enisamium/placebo for clarity. The main text was changed accordingly. We also clarified that enisamium iodide was used in the clinical trial.

