## Supplementary material for "Enisamium is an inhibitor of the SARS-CoV-2 RNA polymerase and shows improvement of recovery in COVID-19 patients in an interim analysis of a clinical trial": Consort checklist

**Items to include when reporting a randomized trial in a journal or conference abstract**

| **Item** | **Description** | **Reported on line number** |
| --- | --- | --- |
| Title | Identification of the study as clinical trial | 1-2 |
| Authors * | Contact details for the corresponding author | 29-30 |
| Trial design | Description of the trial design | 50-53; 500-512 |
| Methods |  |  |
| Participants | Eligibility criteria for participants and the settings where the data were collected | 500-506 |
| Interventions | Interventions intended for each group | 515-520 |
| Objective | Specific objective | 220-230 |
| Outcome | Defined primary outcome | 231-248 |
| Randomization | How participants were allocated to interventions | 234, 500 |
| Blinding (masking) | Whether or not participants, care givers, and those assessing the outcomes were blinded to group assignment | 500 |
| Results |  |  |
| Numbers randomized | Number of participants randomized | 234 |
| Recruitment | Description of recruitment criteria | 500-506 |
| Numbers analysed | Number of participants analysed | 231-240 |
| Outcome | For the primary outcome Kaplan-Meier plots are provided in figure 3. | 231-248 |
| Harms | Important adverse events or side effects | 541-545 |
| Conclusions | General interpretation of the results | 249-252 |
| Trial registration | Registration number and name of trial register | 511-512 |
| Funding | Source of funding | 566-568 |

**this item is specific to conference abstracts*
