## Supplemental figures and tables for "Enisamium is an inhibitor of the SARS-CoV-2 RNA polymerase and shows improvement of recovery in COVID-19 patients in an interim analysis of a clinical trial"

**Contents**:

Supplemental figures: 4

Supplemental tables: 4

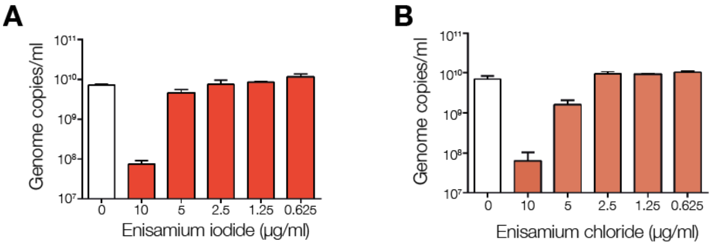


**Figure S1.** Effect of enisamium iodide (**A**) or enisamium chloride (**B**) on SARS-CoV-2 infected Caco-2 cells.

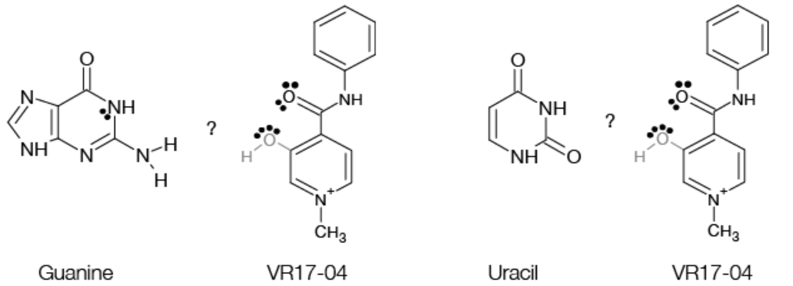


**Figure S2.** VR17-04 is likely not able to form Watson-Crick hydrogen bonds with guanine or uracil.

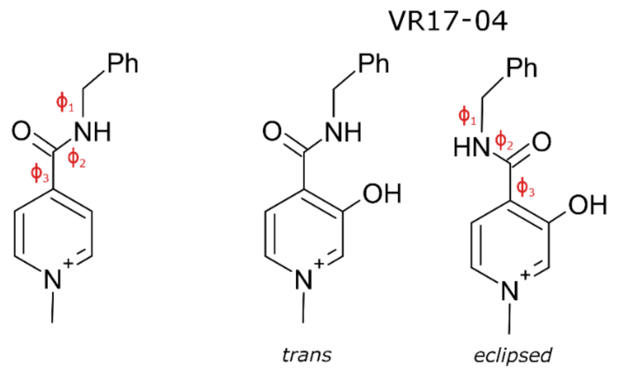


**Figure S3.** Schematic showing enisamium (left) and VR17-04 (middle, right). For both molecules, the $\phi_1$, $\phi_2$, $\phi_3$ dihedral angles are indicated. For VR17-04, the *trans* and *eclipsed* conformations that VR17-04 present at $\phi_3$ are shown.

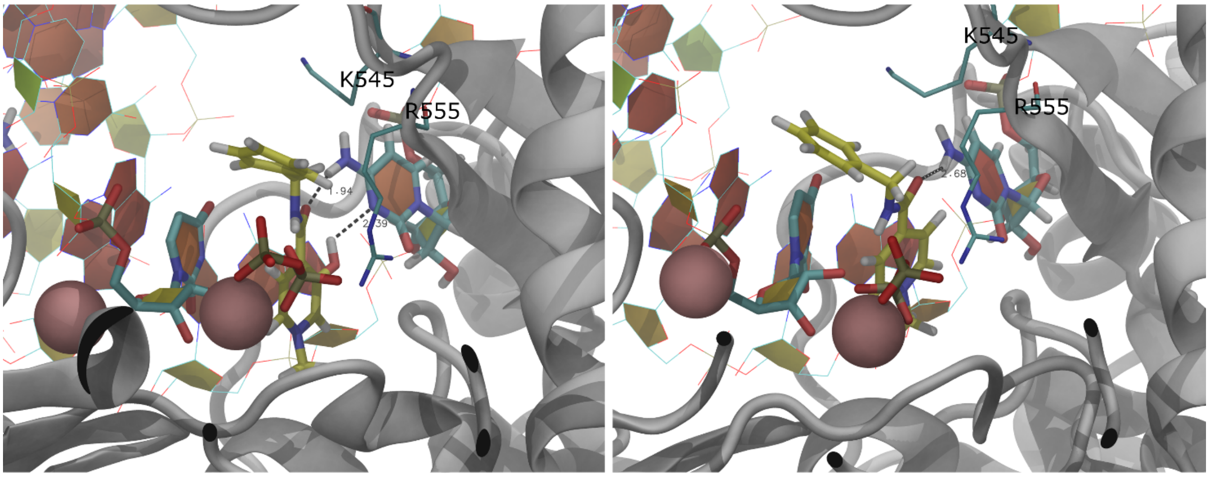


**Figure S4.** Binding of VR17-04 to cytosine (Cyt) in the nsp12/7/8 complex. Snapshots of the (left) VR17-04-nsp12/7/8(Cyt) and (right) enisamium-nsp12/7/8(Cyt) complexes at simulation time 50 and 24 ns. Putative hydrogen bonds and distances between cytosine and the docked enisamium and VR17-04 are shown. The nsp12 protein is shown in grey ribbon representation; the template RNA and nascent strand RNA are drawn in wire representation. Colours cyan, red, blue, and white represent carbon, oxygen, nitrogen, and hydrogen atoms, respectively. The inhibitors VR17-04 and enisamium are represented by tubes, with colours yellow, red, blue, and hydrogen, representing carbon, oxygen, nitrogen, and hydrogen atoms, respectively. The pyr moiety is represented in tubes, with colours orange and red, representing phosphor and oxygen, respectively. The two $Mg^{+2}$ ions are represented by pink spheres. Dashed lines indicate the distances (Å) CO---H2N-Cyt and OH---:N-Cyt in VR17-04-nsp12/7/8(Cyt) and enisamium-nsp12/7/8(Cyt) complexes, respectively. Two key residues of the catalytic cavity are drawn by thin tubes and labelled.

**Table S1.** Quantum chemical (DFT B3LYP/6-31G*) geometry optimization of VR17-04 and enisamium. The dihedral angles ($\phi_1$, $\phi_2$, $\phi_3$) that define the conformation of VR17-04 and enisamium are reported. VR17-04 allows two conformations at $\phi_3$ *trans* and *eclipsed*, respectively. The estimated energies ($E^{B3LYP}$) and zero-point energies ($E^{ZPE}$) are reported in hartree. The zero-point corrected energies ($E^{B3LYP}$+ $E^{ZPE}$) are reported in hartree and in kcal mol$^{-1}$ (bold) to underline the energy difference between the two conformations (*trans*, *eclipsed*) in VR17-04 (8.346 kcal mol$^{-1}$).

| **Compound** | **$\phi_1$** | **$\phi_2$** | **$\phi_3$** | **$\phi_3$conf.** | **$E^{B3LYP}$ (Hartree)** | **$E^{ZPE}$ (hartree)** | $E^{B3LYP}$+ $E^{ZPE}$ (hartree) **(kcal mol$^{-1}$)** |
|---|---|---|---|---|---|---|---|
| VR17-04 | 170 | -179 | 34 | *trans* | -801.7853 | 0.2721 | -801.5131 **(-502’951.875)** |
| VR17-04 | 9 | 177 | -3 | *eclipsed* | -801.7977 | 0.2713 | -801.5264 **(-502’960.221)** |
| Enisamium | 23 | 169 | -21 | - | -726.5961 | 0.2673 | -726.3288 **(-455’773.501)** |

**Table S2.** $^1$H chemical shift of the VR17-04 in water at T = 277 K acquired at 500 MHz. The labelled resonances are reported on the NOE spectra in Fig. 2C.

| $^1$H resonance | HN | H2’ | H6’ | H5’ | Ph | $CH_2$ | $CH_3$ |
|---|---|---|---|---|---|---|---|
| δ ppm | 10.45 | 8.29 | 8.08 | 8.13 | 7.42/7.36 | 4.66 | 4.28 |

**Table S3.** Structural characterization of the interaction (Watson-Crick base pair) between VR17-04-cytosine (Cyt), enisamium-(Cyt), and VR17-04-adenine (Ade), described by average distances ⟨d⟩ between hydrogen bond donor and acceptor groups, and average values of the improper dihedral ⟨Dihed⟩ describing the coplanarity condition. The errors on the mean values are smaller than the last decimal digit. The selected distances and dihedrals are defined by the atoms in bold. The average interval is reported.

| **Complex** | **D** | **⟨*d*⟩ (Å)** | **ϕ** | **⟨ϕ⟩(°)** | **Avg Interval (ns)** |
|---|---|---|---|---|---|
| VR17-04-RdRp(C) | C**O**---**H**$_2$N-Cyt, O**H**---**:N**-Cyt | 1.9, 2.4 | C**O**-H**N**-**N:**-H**O** | 13 | 30-50 |
| Enisamium-RdRp(C) | C**O**---**H**$_2$N-Cyt | 3.3 | C**O**-H**N**-**N:**-**H**C | 20 | 30-50 |
| VR17-04-RdRp(A) | C**O**---**H**$_2$N-Ade, O**H**---**:N**-Ade | 1.9, 2.9 | C**O**-H**N**-**N:**-H**O** | 11 | 20-40 |

**Table S4.** Poisson Boltzmann free energy of binding $\langle \Delta G_{PB}^{Bind} \rangle$ estimated as average value using the MMPBSA methods; the corresponding error on the mean is reported in brackets. The time interval for the average estimation is reported.

| **Inhibitor** | **Complex** | **Average MD interval (ns)** | $\langle \Delta G_{PB}^{Bind} \rangle$ **(kcal mol$^{-1}$)** |
|---|---|---|---|
| VR17-04 | VR17-04-RdRp(Cyt) | [46, 50] | -19.8(4) |
| Enisamium | Enisamium-RdRp(Cyt) | [46, 50] | 43.6(5) |
| VR17-04 | VR17-04-RdRp(Ade) | [28, 32] | -14.8(5) |
